# Evaluation of the diagnostic value of YiDiXie™-SS, YiDiXie™-HS and YiDiXie™-D in uroepithelial carcinoma

**DOI:** 10.1101/2024.08.08.24311656

**Authors:** Huimei Zhou, Shengjie Lin, Yutong Wu, Chen Sun, Xutai Li, Zhenjian Ge, Wenkang Chen, Yingqi Li, Pengwu Zhang, Wuping Wang, Siwei Chen, Wei Li, Yong Xia, Lingzhi Tao, Yongqing Lai

## Abstract

**Background:** Uroepithelial carcinoma is a serious threat to human health and causes heavy economic burden. Enhanced CT is widely used in screening or preliminary diagnosis of uroepithelial tumors. However, false-positive results on enhanced CT can lead to misdiagnosis and incorrect endoscopy, while false-negative results on enhanced CT can lead to missed diagnosis and delayed treatment. There is an urgent need to find convenient, cost-effective and non-invasive diagnostic methods to reduce the false-negative and false-positive rates of enhanced CT in uroepithelial tumors. The aim of this study was to evaluate the diagnostic value of YiDiXie ™ -SS, YiDiXie ™ -HS and YiDiXie ™ -D in uroepithelial carcinoma.

**Patients and methods:** 319 subjects (malignant group, n=240; benign group, n=79) were finally included in this study. Remaining serum samples from the subjects were collected and tested by applying the YiDiXie™ all-cancer detection kit to evaluate the sensitivity and specificity of YiDiXie™-SS and YiDiXie™-HS.

**Results:** The sensitivity of YiDiXie™ SS was 95.8% (95% CI: 92.5% - 97.7%) and its specificity was 64.6% (95% CI: 53.6% - 74.2%). This means that YiDiXie ™ -SS has an extremely high sensitivity and relatively high specificity in urothelial tumors.YiDiXie™-HS has a sensitivity of 85.8% (95% CI: 80.9% - 89.7%) and a specificity of 84.8% (95% CI: 75.3% - 91.1%). This means that YiDiXie™-HS has high sensitivity and specificity in urothelial tumors.YiDiXie™-D has a sensitivity of 73.3% (95% CI: 67.4% - 78.5%) and a specificity of 92.4% (95% CI: 84.4% - 96.5%). This means that YiDiXie ™ -D has relatively high sensitivity and very high specificity in urothelial tumors. The sensitivity of YiDiXie™-SS in enhanced CT-positive patients was 96.3% (95% CI: 96.3% - 98.3%)and its specificity was 64.3% (95% CI: 38.8% - 83.7%). This means that the application of YiDiXie™-SS reduces the false-positive rate of urological enhanced CT by 64.3% (95% CI: 38.8% - 83.7%) with essentially no increase in malignancy leakage. The sensitivity of YiDiXie™-HS in enhanced CT-negative patients was 85.5% (95% CI: 75.9% - 91.7%)and its specificity was 84.6% (95% CI: 73.9% - 91.4%). This means that the application of YiDiXie™-HS reduces the false-negative rate of urological enhanced CT by 85.5% (95% CI: 75.9% - 91.7%).YiDiXie™-D had a sensitivity of 75.6% (95% CI: 68.5% - 81.5%) and a specificity of 92.9% (95% CI: 68.5% - 99.6%) in patients with enhanced CT positivity. This means that YiDiXie ™ -D reduced the rate of false positives in enhanced CT by 92.9% (95% CI: 87.8% - 96.0%). YiDiXie ™ -D has a sensitivity of 68.4% (95% CI: 57.3% - 77.8%) and a specificity of 92.3% (95% CI: 83.2% - 96.7%) in patients with enhanced CT negativity. This means that YiDiXie™-D reduces the false-negative rate of enhanced CT by 68.4% (95% CI: 57.3% - 77.8%) while maintaining high specificity.

**Conclusion:** YiDiXie™-SS has extremely high sensitivity and relatively high specificity in urological tumors. YiDiXie™-HS has high sensitivity and high specificity in urological tumors. YiDiXie™-D has relatively high sensitivity and extremely high specificity in urological tumors. YiDiXie ™ -SS dramatically reduces urological enhanced CT false-positive rates with essentially no increase in delayed treatment of malignant tumors. YiDiXie™-HS substantially reduces urological enhanced CT false-negative rates.YiDiXie ™ -D substantially reduces urological enhanced CT false-positive rates, or significantly reduces urological enhanced CT false-negative rates while maintaining high specificity. YiDiXie™ tests has important diagnostic value in uroepithelial cancer, and is expected to solve the problems of “high false positive rate” and “high false negative rate” of urological enhanced CT.

**Clinical trial number:** ChiCTR2200066840.

## INTRODUCTION

Uroepithelial carcinoma (UC) is a common malignant tumor that includes carcinoma of renal pelvis, ureter cancer and bladder cancer. Data show that upper uroepithelial carcinoma of the renal pelvis or ureter is relatively rare, with an incidence of about 2/100,000, accounting for only 5-10% of cases^1,2^. However, 90% of uroepithelial cancers are uroepithelial cancers of the bladder, which account for approximately 49.1% of all new cases in the urinary system^3^. There will be approximately 573,000 new cases and 213,000 deaths from bladder cancer only in 2020^4^. There are more men than women, with male incidence and mortality rates of 9.5 and 3.3 cases per 100,000 people, respectively, about four times that of women globally^4^. In 2022, it adds to about 614,000 new cases and 220,000 deaths^5^. Today, treatments for uroepithelial cancer include surgery, endoscopic treatment, bladder irrigation, systemic chemotherapy and radiotherapy, but the predominant approach is surgery. Surgical procedures may include radical resection and partial resection^6,7^. Five-year disease-specific survival in patients with upper urothelial carcinoma is relatively low, as 60% of patients have invasive carcinoma at the time of diagnosis and approximately 9% have metastases^7,8^. And for bladder cancer, patients with non-muscle-invasive bladder cancer (NMIBC) have a five-year survival rate of up to 90 while muscle-invasive bladder cancer (MIBC) patients have a 5-year overall survival rate of only about 50%. The 5-year overall survival rate for metastatic bladder cancer patients is even lower at 5%^9,10^. In addition, the per capita cost of diagnosis to death for patients with bladder cancer is the highest of all malignancies due to the frequent procedures required for disease surveillance and treatment^11^. Therefore, uroepithelial carcinoma is a serious threat to human health and a heavy economic burden.

Enhanced CT is widely used in the diagnosis of uroepithelial tumors. On the one hand, urinary tract enhanced CT can produce a large number of false-positive results. When urological enhanced CT is positive, patients usually take endoscopy and biopsy pathology. False-positive results of urological enhanced CT mean that benign diseases are misdiagnosed as malignant tumors, and patients will have to bear unnecessary mental pain, expensive examination costs, physical injuries, and other adverse consequences. Therefore, there is an urgent need to find a convenient, economical and non-invasive diagnostic method to reduce the false-positive rate of urological enhanced CT.

On the other hand, urological enhanced CT can produce a large number of false-negative results. When urological enhanced CT is negative, patients usually take observation and regular follow-up. False-negative results of urological enhanced CT imply misdiagnosis of malignant tumors as benign diseases, which will likely lead to delayed treatment, progression of malignant tumors, and possibly even development of advanced stages. Patients will thus have to bear the adverse consequences of poor prognosis, high treatment costs, poor quality of life, and short survival. Therefore, there is an urgent need to find a convenient, economical and noninvasive diagnostic method to reduce the false-negative rate of urological enhanced CT.

Based on the detection of novel tumor markers of miRNA in serum, Shenzhen KeRuiDa Health Technology Co., Ltd. has developed an in vitro diagnostic test, YiDiXie ™ all-cancer tests (hereinafter referred to as YiDiXie ™ tests), which can detect multiple types of cancers with only 200 microliters of whole blood or 100 microliters of serum each time^12^. YiDiXie™ tests consists of three different tests, YiDiXie ™ -SS, YiDiXie ™ -HS and YiDiXie™-D^12^.

The purpose of this study was to evaluate the diagnostic value of YiDiXie™-SS, YiDiXie™-HS and YiDiXie™-D in uroepithelial carcinoma.

## PATIENTS AND METHODS

### Study design

This work is part of the sub-study “Evaluating the diagnostic value of YiDiXie™ tests in multiple tumors” of the SZ-PILOT study (ChiCTR2200066840).

The SZ-PILOT study (ChiCTR2200066840) is an observational, prospective, single-center investigation. All participants in the study were required to complete an informed consent form at the time of admission or physical examination for the donation of residual samples. The study collected the remaining 0.5 ml of serum sample for analysis.

It was a blinded trial, with the laboratory staff conducting the YiDiXie test and the KeRuiDa laboratory technicians calculating the results being unaware of the clinical information about the individuals. Similarly, the YiDiXie test results were unknown to the clinical specialists who evaluated the participants’ clinical data.

The study was approved by the Ethics Committee of Peking University Shenzhen Hospital and was conducted in accordance with the International Conference on Harmonization for “Good clinical practice guidelines” and the Declaration of Helsinki.

### Participants

This study included participants who had a positive ultrasonography examination for urinary tract. The two groups of subjects were enrolled independently, and each subject who met the inclusion criteria was added sequentially.

Initially, inpatients with “suspected (solid or haematological) malignancy” who had provided general informed consent for the donation of the remaining samples were included in the study. The study classified subjects into two groups based on their postoperative pathological diagnosis: those with a diagnosis of “malignant tumor” and those with a diagnosis of “benign tumor”. Some of the samples from the malignant group were used in our previous work^12^.

Subjects with unclear pathology findings were excluded from the study. For information on enrollment and exclusion, please refer to our previous work^12^.

### Sample collection, processing

The serum samples utilized in this investigation were derived from surplus serum following consultation and treatment, without additional blood donation. Around 0.5 ml aliquots of the residual serum from the participants were procured in the clinical laboratory and preserved at -80°C for subsequent employment in the YiDiXie test.

### The YiDiXie test

The YiDiXie test is executed using the YiDiXie all-cancer detection kit, which is developed and manufactured by Shenzhen KeRuiDa Health Technology Co..^12^ It utilizes fluorescent PCR technology as its in-vitro diagnostic system. To ascertain the presence of cancer in the subjects, it gauges the expression levels of numerous miRNA biomarkers in the serum. The predefined thresholds for each miRNA biomarker ensures high specificity for each miRNA marker. The amalgamation of independent assays into a parallel trial format culminates in a substantial enhancement in sensitivity and high specificity for a wide range of cancers.^12^

YiDiXie ™ tests consists of three distinctly different tests: YiDiXie™-Highly Sensitive (YiDiXie™ -HS), YiDiXie ™ -Super Sensitive(YiDiXie ™ -SS) and YiDiXie ™ -Diagnosis (YiDiXie ™ -D).^12^ YiDiXie ™ -HS has been developed with sensitivity and specificity in mind.^12^ YiDiXie ™ -SS significantly increased the number of miRNA tests to achieve extremely high sensitivity for all clinical stages of all malignancy types.^12^ YiDiXie ™ -D dramatically increases the diagnostic threshold of individual miRNA tests to achieve very high specificity (very low false diagnosis rate) for all malignancy types.^12^

Perform YiDiXie ™ tests according to the instructions provided by the YiDiXie ™ all-cancer detection kit. The detailed procedure can be found in our previous work^12^.

The laboratory technicians of Shenzhen KeRuiDa Health Technology Co., Ltd examined the raw test results and determined that YiDiXie™ tests had either “positive” or “negative” results^12^.

### Diagnosis of Enhanced CT

The diagnostic conclusion of urological enhanced CT is considered “positive” or “negative”. The test result is considered “positive” if the diagnosis is positive, more positive, or favors a malignant tumor. If the diagnosis is positive, more positive, or favors a benign tumor, or if the diagnosis is ambiguous, the test result is considered “negative”.

### Extraction of clinical data

The subjects’ inpatient medical records or physical examination reports were used to extract clinical, pathological, laboratory, and imaging data for this study. The AJCC staging manual (7th or 8th edition) was used by trained clinicians to complete clinical staging^13-14^.

### Statistical analyses

Descriptive statistics were reported for demographic and baseline characteristics. For categorical variables, the number and percentage of subjects in each category were calculated; For continuous variables, the total number of subjects (n), mean, standard deviation (SD) or standard error (SE), median, first quartile (Q1), third quartile (Q3), minimum, and maximum values were calculated. The Wilson (score) method was used to calculate 95% confidence intervals (CIs) for multiple indicators.

## RESULTS

### Participant disposition

319 subjects (malignant group, n=240; benign group, n=79) were finally included in this study. The demographic and clinical characteristics of the 319 study subjects are presented in Table 1.

**Table 1.**
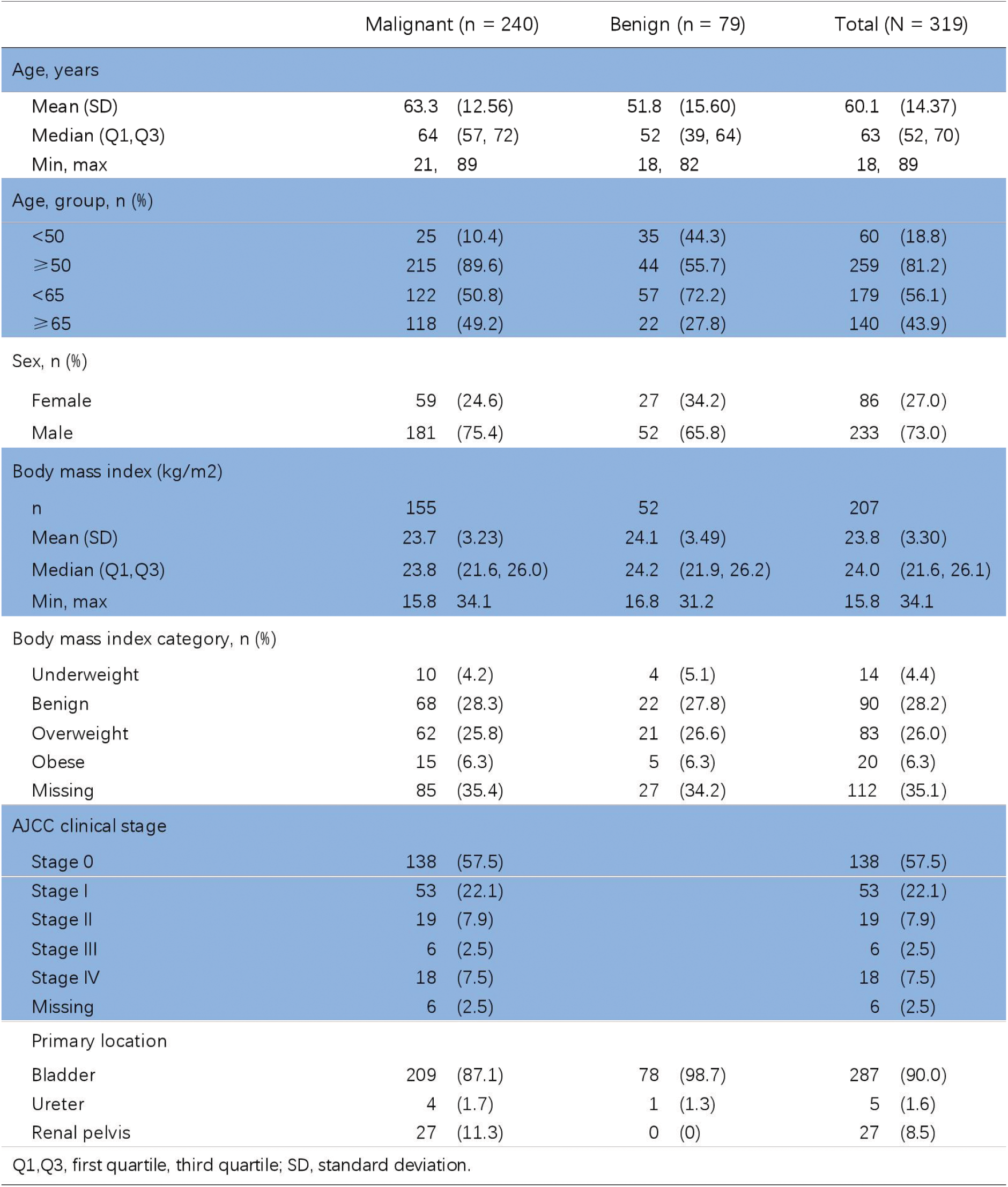
Participants’ demographic and clinical manifestation.

The two groups of study subjects were comparable in terms of demographic and clinical characteristics (Table 1). The mean (standard deviation) age was 60.1(14.37) years and 27.0%(86/319) were female.

### Diagnostic performance of YiDiXie™-SS

As shown in Table 2, the sensitivity of YiDiXie™ -SS was 95.8% (95% CI: 92.5% - 97.7%) and its specificity was 64.6% (95% CI: 53.6% - 74.2%). This means that YiDiXie ™ -SS has very high sensitivity and relatively high specificity in urothelial tumors.

**Table 2.** Performance of YiDiXie™ –SS.

| Table 2. Performance of YiDiXie™-SS |  |  |  |
| --- | --- | --- | --- |
|  | Malignant | Benign | Total |
|  | 240 | 79 | 319 |
| Positive | 230 | 28 | 258 |
| Negative | 10 | 51 | 61 |
| SEN = 230/240 = 95.8% (92.5% - 97.7%) |  | SPE = 51/79 = 64.6% (53.6% - 74.2%) |  |

SEN, Sensitivity. SPE, Specificity. Two-sided 95% Wilson confidence intervals were calculated.

### Diagnostic performance of YiDiXie™-HS

As shown in Table 3, the sensitivity of YiDiXie™ -HS was 85.8% (95% CI: 80.9% - 89.7%) and its specificity was 84.8% (95% CI: 75.3% - 91.1%). This means that YiDiXie ™ -HS has high sensitivity and high specificity in urinary tract tumors.

**Table 3.** Performance of YiDiXie™ –HS.

| Table 3. Performance of YiDiXie™-HS |  |  |  |
| --- | --- | --- | --- |
|  | Malignant | Benign | Total |
|  | 240 | 79 | 319 |
| Positive | 206 | 12 | 218 |
| Negative | 34 | 67 | 101 |
| SEN = 206/240 = 85.8% (80.9% - 89.7%) |  | SPE = 67/79 = 84.8% (75.3% - 91.1%) |  |

SEN, Sensitivity. SPE, Specificity. Two-sided 95% Wilson confidence intervals were calculated.

### Diagnostic performance of YiDiXie™-D

As shown in Table 4, the sensitivity of YiDiXie™ -D was 73.3% (95% CI: 67.4% - 78.5%) and its specificity was 92.4% (95% CI: 84.4% - 96.5%). This means that YiDiXie ™ -D has high sensitivity and very high specificity in urothelial tumors.

**Table 4.** Performance of YiDiXie™ –D.

|  | Malignant | Benign | Total |
| --- | --- | --- | --- |
|  | 240 | 79 | 319 |
| Positive | 176 | 6 | 182 |
| Negative | 64 | 73 | 137 |
| SEN = 176/240 = 73.3% (67.4% - 78.5%) |  |  | SPE = 73/79 = 92.4% (84.4% - 96.5%) |

SEN, Sensitivity. SPE, Specificity. Two-sided 95% Wilson confidence intervals were calculated.

### Diagnostic performance of enhanced CT

As shown in Table 5, the sensitivity of enhanced CT was 68.3% (95% CI: 62.2% - 73.9%)and its specificity was 82.3% (95% CI: 72.4% - 89.1%).

**Table 5.** Performance of different Items.

| Items | Total N | Positive | Sensitivity % (95% CI)<sup>a</sup> | Total N | Negative | Specificity % (95% CI)<sup>b</sup> |
| --- | --- | --- | --- | --- | --- | --- |
| CT | 240 | 164 | 68.3% (62.2% - 73.9%) | 79 | 65 | 82.3% (72.4% - 89.1%) |
| YiDiXie™-SS in CT-positive | 164 | 158 | 96.3% (92.2% - 98.3%) | 14 | 9 | 64.3% (38.8% - 83.7%) |
| YiDiXie™-HS in CT-negative | 76 | 65 | 85.5% (75.9% - 91.7%) | 65 | 55 | 84.6% (73.9% - 91.4%) |
| YiDiXie™-D in CT-positive | 164 | 124 | 75.6% (68.5% - 81.5%) | 14 | 13 | 92.9% (68.5% - 99.6%) |
| YiDiXie™-D in CT-negative | 76 | 52 | 68.4% (57.3% - 77.8%) | 65 | 60 | 92.3% (83.2% - 96.7%) |

CI, confidence interval. <sup>a,b</sup> Two-sided 95% Wilson confidence intervals were calculated.

### Diagnostic Performance of YiDiXie™-SS in enhanced CT-positive patients

To address the challenge of high false-positive rate of urological enhanced CT, YiDiXie ™ -SS was applied to enhanced CT-positive patients.

As shown in Table 5, the sensitivity of YiDiXie™ -SS in enhanced CT-positive patients was 96.3% (95% CI: 96.3% - 98.3%)and its specificity was 64.3%

(95% CI: 38.8% - 83.7%). This means that the application of YiDiXie ™ -SS reduces the false-positive rate of urological enhanced CT by 64.3% (95% CI: 38.8% - 83.7%) with essentially no increase in malignancy leakage.

### Diagnostic Performance of YiDiXie™-HS in enhanced CT-negative patients

To address the challenge of high false-negative rate of urological enhanced CT, YiDiXie™-HS was applied to enhanced CT-negative patients.

As shown in Table 5, the sensitivity of YiDiXie™ -HS in enhanced CT-negative patients was 85.5% (95% CI: 75.9% - 91.7%)and its specificity was 84.6% (95% CI: 73.9% - 91.4%). This means that the application of YiDiXie ™ -HS reduces the false-negative rate of urological enhanced CT by 85.5% (95% CI: 75.9% - 91.7%).

### Diagnostic Performance of YiDiXie™-D in enhanced CT-positive patients

In order to further minimize false-positive results in patients with enhanced CT positivity, the more specific YiDiXie™-D was therefore applied to such patients.

As shown in Table 5, YiDiXie ™ -D had a sensitivity of 75.6% (95% CI: 68.5% - 81.5%) and its specificity was 92.9% (95% CI: 68.5% - 99.6%) in enhanced CT positive patients. This means that YiDiXie ™ -D reduces the false-positive rate of enhanced CT by 92.9% (95% CI: 87.8% - 96.0%).

### Diagnostic Performance of YiDiXie™-HS in enhanced CT-negative patients

In order to further minimize false-positive results in enhanced CT-negative patients, the more specific YiDiXie™-D was therefore applied to such patients.

As shown in Table 5, YiDiXie ™ -D had a sensitivity of 68.4% (95% CI: 57.3% - 77.8%) and its specificity was 92.3% (95% CI: 83.2% - 96.7%) in patients with enhanced CT negativity. This means that YiDiXie™-D reduces the false-negative rate of enhanced CT by 68.4% (95% CI: 57.3% - 77.8%) while maintaining high specificity.

## DISCUSSION

### Clinical significance of YiDiXie™-SS in enhanced CT-positive patients

YiDiXie ™ tests consists of 3 tests with very different characteristics: YiDiXie™-SS, YiDiXie™-HS and YiDiXie ™ -D^12^. Among them, YiDiXie ™ -HS combines high sensitivity and high specificity^12^. YiDiXie ™ -SS has very high sensitivity for all malignant tumor types, but slightly lower specificity^12^. YiDiXie™-D has very high specificity for all malignant tumor types, but lower sensitivity^12^.

For urological enhanced CT-positive patients, the sensitivity and specificity of further diagnostic methods are important. The trade-off between sensitivity and specificity is essentially a trade-off between the “danger of underdiagnosis of malignant tumors” and the “danger of misdiagnosis of benign diseases”. In general, a positive urological enhanced CT is usually followed by endoscopy and biopsy rather than radical surgery. Therefore, a false-positive urological enhanced CT does not lead to serious consequences such as major surgical trauma, organ removal, or loss of function. Thus, the “risk of malignant diagnosis” is much higher than the “risk of benign disease misdiagnosis” in patients with a positive urological enhanced CT. Therefore, YiDiXie ™ -SS, which has very high sensitivity but slightly lower specificity, was chosen to reduce the false-positive rate of urological enhanced CT.

As shown in Table 5, YiDiXie ™ -SS had a sensitivity of 96.3% (95% CI: 96.3% - 98.3%) and a specificity of 64.3% (95% CI: 38.8% - 83.7%) in patients with positive urological enhanced CT. The above results indicate that YiDiXie™-SS reduces the magnitude of urological enhanced CT false positives by 64.3% (95% CI: 38.8% - 83.7%), while maintaining a sensitivity close to 100%.

This means that YiDiXie ™ -SS dramatically reduces the probability of unnecessary endoscopy with biopsy in patients with benign urological diseases, without essentially increasing the number of malignant tumors missed. In other words, YiDiXie ™ -SS substantially reduces the mental anguish, expensive examination costs, examination injuries, and other adverse consequences for urological enhanced CT false-positive patients with essentially no increase in delayed treatment of malignant tumors. Therefore, YiDiXie ™ -SS well meets the clinical needs and has important clinical significance and wide application prospects.

### Clinical significance of YiDiXie™-HS in enhanced CT-negative patients

For urological enhanced CT-negative patients, the sensitivity and specificity of further diagnostic methods are important. The trade-off between both the sensitivity and specificity is essentially a trade-off between the “danger of malignant tumor underdiagnosis” and the “danger of benign disease misdiagnosis”. Higher false-negative rates mean that more malignant tumors are underdiagnosed, leading to delays in treatment, progression of malignant tumors, and even advanced stages. As a result, patients will have to bear the adverse consequences of poor prognosis, short survival, poor quality of life, and high treatment costs. Higher false-positive rate means more misdiagnosis of benign diseases, which will lead to unnecessary expensive and invasive endoscopy and biopsy. As a result, patients have to bear the consequences of mental suffering, expensive tests, and injuries. Therefore, YiDiXie ™-HS with its high sensitivity and specificity was chosen to reduce the false negative rate of urological enhanced CT.

As shown in Table 5, YiDiXie ™ -HS had a sensitivity of 85.5% (95% CI: 75.9% - 91.7%) and a specificity of 84.6% (95% CI: 73.9% - 91.4%) in patients with urological enhanced CT negativity. The above results indicate that YiDiXie ™ -HS reduced false negatives in urological enhanced CT by 85.5% (95% CI: 75.9% - 91.7%).

The above results imply that YiDiXie ™ -HS significantly reduces the probability of malignant tumors being missed by urological enhanced CT. In other words, YiDiXie™-HS substantially reduces the poor prognosis, high treatment cost, poor quality of life, and short survival of patients with missed diagnosis of urological enhanced CT. Therefore, YiDiXie™-HS well meets the clinical needs and has important clinical significance and wide application prospects.

### Clinical significance of YiDiXie™-D

For patients with urologic tumors, YiDiXie™-D, which has relatively high sensitivity and very high specificity, can be used to further reduce the false-positive rate of Urologic enhanced CT or to significantly reduce its false-negative rate while maintaining a high level of specificity.

As shown in Table 5, YiDiXie ™ -D had a sensitivity of 75.6% (95% CI: 68.5% - 81.5%) and a specificity of 92.9% (95% CI: 68.5% - 99.6%) in patients with positive enhanced CT, and 68.4% (95% CI: 57.3% - 77.8%) in patients with negative enhanced CT. 77.8%) and its specificity was 92.3% (95% CI: 83.2% - 96.7%) in patients with negative enhanced CT. These results suggest that YiDiXie™ -D reduces the false-positive rate of enhanced CT by 92.9% (95% CI: 87.8% - 96.0%) or reduces the false-negative rate of enhanced CT by 68.4% (95% CI: 57.3% - 77.8%) while maintaining a high specificity.

The above results imply that YiDiXie ™ -D further reduces the risk of incorrect endoscopy for urothelial tumors. Therefore, YiDiXie™-D meets the clinical needs well and has important clinical significance and wide application prospects.

### YiDiXie™ tests is expected to address 2 challenges in Uroepithelial Carcinoma

First, YiDiXie™-SS can greatly relieve clinicians of non-essential workloads and facilitate timely diagnosis and treatment of malignant tumor cases that would otherwise be delayed. with a positive urological enhanced CT, endoscopy and biopsy are commonly performed on this patient. The timely completion of these endoscopies and biopsies is directly dependent on the number of clinicians. In many parts of the world, appointments are made for months or even more than a year. This inevitably delays the treatment of malignancy cases among them, and therefore it is not uncommon for urological enhanced CT positive patients waiting for endoscopy with biopsy to have malignancy progression or even distant metastasis. As shown in the results, YiDiXie™-SS reduced the false-positive rate of urological enhanced CT by 64.3% (95% CI: 38.8% - 83.7%) with essentially no increase in malignancy leakage. As a result, YiDiXie ™-SS can significantly relieve clinicians of unnecessary workloads and facilitate timely diagnosis and treatment of uroepithelial cancers or other diseases that would otherwise be delayed.

Second, YiDiXie™-HS greatly reduces the risk of underdiagnosis of uroepithelial cancer. when urological enhanced CT is negative, the possibility of uroepithelial cancer is usually ruled out for the time being. The high rate of false-negative urological enhanced CT results in delayed treatment for a large number of patients with uroepithelial cancer. As shown in the results, YiDiXie ™ -HS reduced the false negative rate of urological enhanced CT by 85.5% (95% CI: 75.9% - 91.7%). As a result, YiDiXie ™ -HS significantly reduced the probability of missed malignancies due to false-negative urological enhanced CT, facilitating timely diagnosis and treatment of patients with uroepithelial cancers that would otherwise be delayed.

Third, YiDiXie ™ -D is expected to further address the challenges of “high false positive rate” and “high false negative rate”. As shown in Table 5, YiDiXie ™ -D reduced the false-positive rate of enhanced CT by 92.9% (95% CI: 87.8% - 96.0%) or the false-negative rate of enhanced CT by 68.4% (95% CI: 57.3% - 77.8%) while maintaining a high specificity. As a result, YiDiXie™-D further reduces the risk of incorrect endoscopy for urothelial tumors.

Final, the YiDiXie ™ -SS enables “just-in-time” diagnosis of patients with uroepithelial carcinoma. For one, YiDiXie ™ tests requires only trace amounts of blood, allowing patients to complete the diagnostic process non-invasively at home. Only 20 microliters of serum are required to complete a single YiDiXie ™ tests, which is equivalent to approximately 1 drop of whole blood (1 drop of whole blood is approximately 50 microliters, which yields 20-25 microliters of serum)^12^. Taking into account the pre-test sample quality assessment test and 2-3 repetitions, 0.2 ml of whole blood is adequate to complete YiDiXie™ tests^12^. 0.2 ml of finger blood can be collected at home by the average patient with a finger blood collection needle, instead of requiring venous blood collection by medical personnel^12^. Patients can complete the entire diagnostic process non-invasively without having to leave their homes^12^.

For the other, the diagnostic capacity of YiDiXie™-SS is nearly limitless. Figure 1 shows the basic flowchart of YiDiXie ™ tests, from which we can see that YiDiXie ™ tests not only requires no physicians or medical equipment, but also requires no medical personnel to collect blood^12^. Therefore, YYiDiXie ™ tests is completely independent of clinicians and medical facilities, and its testing capacity is nearly unlimited^12^. As a result, YiDiXie™ tests enables “just-in-time” diagnosis of patients with uroepithelial carcinoma with no need for patients to wait anxiously for an appointment.

**Figure 1.**
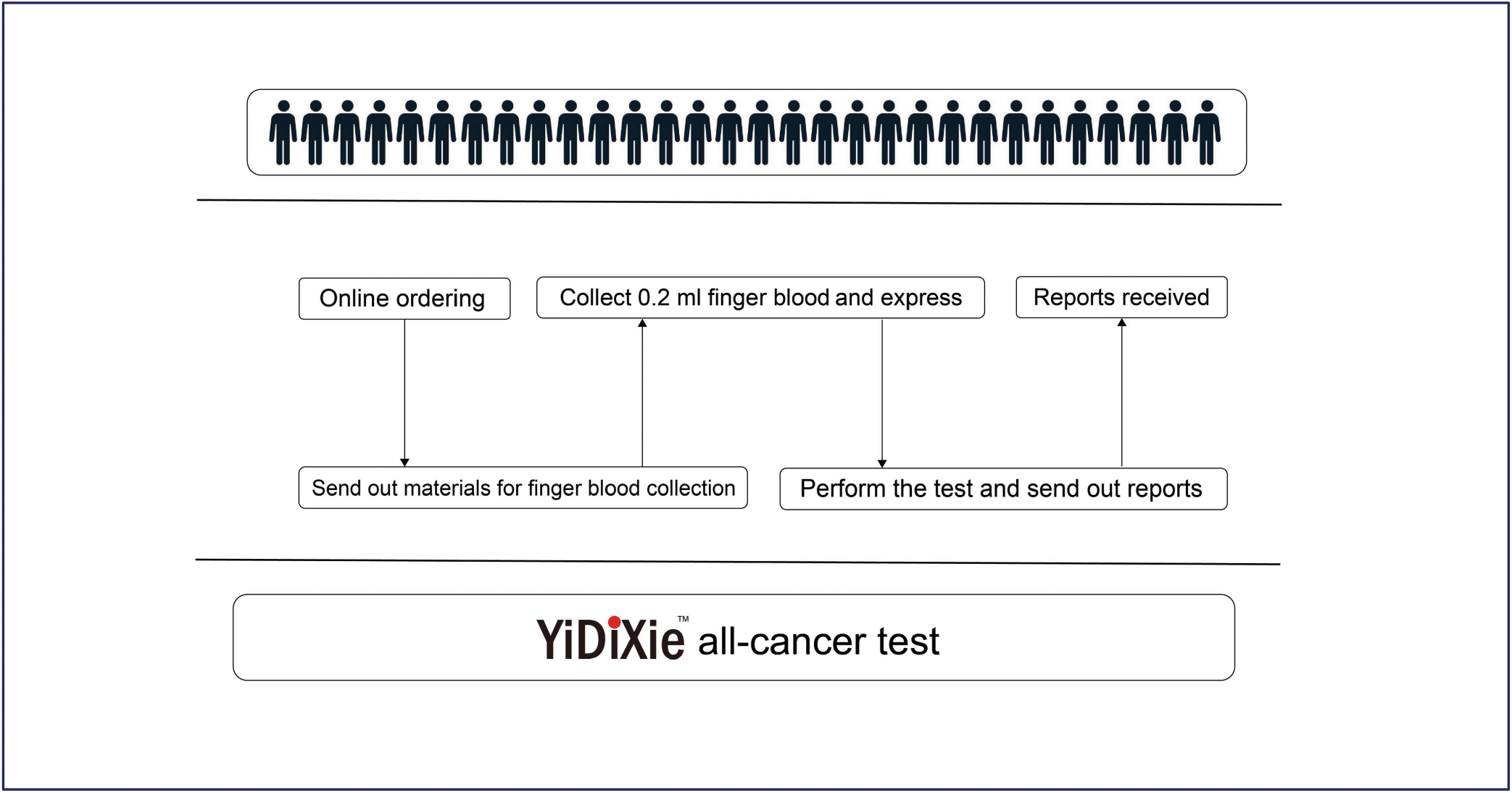
Basic flowchart of YiDiXie™ tests.

In short, YiDiXie ™ tests has an important diagnostic value in uroepithelial carcinoma, and is expected to solve the problems of “high false-positive rate” and “high false-negative rate” of urological enhanced CT in uroepithelial carcinoma.

### Limitations of the study

Firstly, the number of cases in this study was small and future clinical studies with larger sample sizes are needed for further evaluation.

Secondly, this study was a malignant tumor case-benign tumor control study in inpatients, and future cohort studies of patients with positive uroepithelial ultrasound are needed for further assessment.

Finally, this study was a single-centre study, which may have led to some degree of bias in the results of this study. Future multi-centre studies are needed to further assess this.

## CONCLUSION

YiDiXie™-SS has extremely high sensitivity and relatively high specificity in urological tumors. YiDiXie ™ -HS has high sensitivity and high specificity in urological tumors. YiDiXie ™ -D has relatively high sensitivity and extremely high specificity in urological tumors. YiDiXie ™ -SS dramatically reduces urological enhanced CT false-positive rates with essentially no increase in delayed treatment of malignant tumors. YiDiXie ™ -HS substantially reduces urological enhanced CT false-negative rates.YiDiXie ™ -D substantially reduces urological enhanced CT false-positive rates, or significantly reduces urological enhanced CT false-negative rates while maintaining high specificity. YiDiXie™ tests has important diagnostic value in uroepithelial cancer, and is expected to solve the problems of “high false positive rate” and “ high false negative rate” of urological enhanced CT.

## Data Availability

All data produced in the present study are contained in the manuscript.

## REFERENCES

1. Rouprêt M, Zigeuner R, Palou J, Boehle A, Kaasinen E, Sylvester R, Babjuk M, Oosterlinck W: European guidelines for the diagnosis and management of upper urinary tract urothelial cell carcinomas: 2011 update. European urology 2011, 59(4):584–594.

2. Rouprêt M, Seisen T, Birtle AJ, Capoun O, Compérat EM, Dominguez-Escrig JL, Gürses Andersson I, Liedberg F, Mariappan P, Hugh Mostafid A et al: European Association of Urology Guidelines on Upper Urinary Tract Urothelial Carcinoma: 2023 Update. European urology 2023, 84(1):49–64.

3. Siegel RL, Giaquinto AN, Jemal A: Cancer statistics, 2024. CA Cancer J Clin 2024, 74(1):12–49.

4. Sung H, Ferlay J, Siegel RL, Laversanne M, Soerjomataram I, Jemal A, Bray F: Global Cancer Statistics 2020: GLOBOCAN Estimates of Incidence and Mortality Worldwide for 36 Cancers in 185 Countries. CA Cancer J Clin 2021, 71(3):209–249.

5. Bray F, Laversanne M, Sung H, Ferlay J, Siegel RL, Soerjomataram I, Jemal A: Global cancer statistics 2022: GLOBOCAN estimates of incidence and mortality worldwide for 36 cancers in 185 countries. CA Cancer J Clin 2024, 74(3):229–263.

6. Holzbeierlein J, Bixler BR, Buckley DI, Chang SS, Holmes RS, James AC, Kirkby E, McKiernan JM, Schuckman A: Treatment of Non-Metastatic Muscle-Invasive Bladder Cancer: AUA/ASCO/SUO Guideline (2017; Amended 2020, 2024). The Journal of urology 2024, 212(1):3–10.

7. Coleman JA, Clark PE, Bixler BR, Buckley DI, Chang SS, Chou R, Hoffman-Censits J, Kulkarni GS, Matin SF, Pierorazio PM et al: Diagnosis and Management of Non-Metastatic Upper Tract Urothelial Carcinoma: AUA/SUO Guideline. The Journal of urology 2023, 209(6):1071–1081.

8. Margulis V, Shariat SF, Matin SF, Kamat AM, Zigeuner R, Kikuchi E, Lotan Y, Weizer A, Raman JD, Wood CG: Outcomes of radical nephroureterectomy: a series from the Upper Tract Urothelial Carcinoma Collaboration. Cancer 2009, 115(6):1224–1233.

9. Pakmanesh H, Anvari O, Forey N, Weiderpass E, Malekpourafshar R, Iranpour M, Shahesmaeili A, Ahmadi N, Bazrafshan A, Zendehdel K et al: TERT Promoter Mutations as Simple and Non-Invasive Urinary Biomarkers for the Detection of Urothelial Bladder Cancer in a High-Risk Region. International journal of molecular sciences 2022, 23(22).

10. Gurney H, Clay TD, Oliveira N, Wong S, Tran B, Harris C: Systemic treatment of advanced and metastatic urothelial cancer: The landscape in Australia. Asia-Pacific journal of clinical oncology 2023, 19(6):585–595.

11. Botteman MF, Pashos CL, Redaelli A, Laskin B, Hauser R: The health economics of bladder cancer: a comprehensive review of the published literature. PharmacoEconomics 2003, 21(18):1315–1330.

12. Chen Sun, Chong Lu, Yongjian Zhang, Ling Wang, Zhenjian Ge, Zhenyu Wen, Wenkang Chen, Yingqi Li, Yutong Wu, Shengjie Lin et al: Evaluation of the Multi-Cancer Early Detection (MCED) value of YiDiXie ™ -HS and YiDiXie™-SS. medRxiv 2024: doi: 10.1101/2024.1103.1111.24303683.

13. Edge SB, Compton CC: The American Joint Committee on Cancer: the 7th edition of the AJCC cancer staging manual and the future of TNM. Ann Surg Oncol 2010, 17(6):1471–1474.

14. Amin MB, Greene FL, Edge SB, Compton CC, Gershenwald JE, Brookland RK, Meyer L, Gress DM, Byrd DR, Winchester DP: The Eighth Edition AJCC Cancer Staging Manual: Continuing to build a bridge from a population-based to a more “personalized” approach to cancer staging. CA Cancer J Clin 2017, 67(2):93–99.

